# Test to release from isolation after testing positive for SARS-CoV-2

**DOI:** 10.1101/2022.01.04.21268372

**Authors:** Billy J. Quilty, Juliet R. C. Pulliam, Carl A. B. Pearson

## Abstract

- The rapid spread and high transmissibility of the Omicron variant of SARS-CoV-2 is likely to lead to a significant number of key workers testing positive simultaneously.
- Under a policy of self-isolation after testing positive, this may lead to extreme staffing shortfalls at the same time as e.g. hospital admissions are peaking.
- Using a model of individual infectiousness and testing with lateral flow tests (LFT), we evaluate test-to-release policies against conventional fixed-duration isolation policies in terms of excess days of infectiousness, days saved, and tests used.
- We find that the number of infectious days in the community can be reduced to almost zero by requiring at least 2 consecutive days of negative tests, regardless of the number of days’ wait until testing again after initially testing positive.
- On average, a policy of fewer days’ wait until initiating testing (e.g 3 or 5 days) results in more days saved vs. a 10-day isolation period, but also requires a greater number of tests.
- Due to a lack of specific data on viral load progression, infectivity, and likelihood of testing positive by LFT over the course of an Omicron infection, we assume the same parameters as for pre-Omicron variants and explore the impact of a possible shorter proliferation phase.

---

The Omicron variant of SARS-CoV-2 (Pango lineage B.1.1.529) was first reported in South Africa on November 25, 2021 (1) and subsequently designated a variant of concern by the WHO on November 27, 2021 (2). Incidence trends in South Africa, which were subsequently replicated in the UK and elsewhere, indicate substantial growth advantage against prevailing variants (3). Empirical and theoretical studies indicate this advantage is due to a combination of immune evasion (4–6) and higher transmissibility (7,8). As such, the spread of Omicron is likely to lead to a resurgence in infections and hospitalisation, even in populations with high rates of prior exposure and/or vaccination.

High rates of infection are likely to cause a high proportion of workers to be off-work isolating after a positive test, typically for 10 days (9). Increases in the number of COVID-19 hospitalisations will increase the exposure risk of healthcare workers, who will be required to isolate after testing positive, compounding the pressure on health services. Provided an individual’s infection is mild and they feel able to, it may be possible for them to return to work earlier if they are no longer infectious. Here, we explore the possibility of shortening the 10-day isolation period, with or without the requirement of negative rapid antigen lateral flow tests (LFTs) to release, while monitoring excess days of infectiousness. To evaluate the practicability of this scheme in test-limited settings, we also consider delaying the start of testing-for-release after the initial positive test, and the number of consecutive days of negative results required for release.

## Method

We create sample populations of 10,000 individuals, each with a viral load trajectory defined by a proliferation and clearance phase duration (in days) and peak viral load (in RNA copies/ml) sampled from distributions (assumed to be Normal) reported by Kissler et al. (10) (Figure S1). We then estimate the probability of being infectious (p_inf_) (assumed equal to that of having culturable virus) and testing positive by the “Biotime Innova” LFT (p_pos_) given viral load by fitting a logistic regression model to data from Pickering et al. (11). Using this model, we then define whether an individual is infectious and/or test-positive given their viral load each day over the course of infection as Bernoulli(p_inf_(t)) and Bernoulli(p_pos_(t)) respectively.

We then apply different policy scenarios to evaluate those schemes in terms of days of isolation avoided compared to a 10-day isolation period, the days of infectiousness following release from isolation and the number of tests used per 10,000 individuals. We consider the following policy dimensions: 3, 5, or 7 days’ wait until initiating (or resuming) testing and 1, 2, or 3 days of consecutive negative tests required for release. We also compare against a policy of shorter isolation without testing to release.

We used the viral load trajectories for vaccinated and unvaccinated individuals from Kissler et al., and assume individuals initially test daily each morning (as is policy for contacts of cases in the UK as of December 2021 (12)), begin isolating from their first positive test before entering into the specified policy scenario. We do not consider isolation due to symptom onset in this analysis. We assume an upper bound for the isolation period of 10 days regardless of test status. Possible pathways are shown in Figure S2. For sensitivity analysis, we explore the implications of a possibly shorter incubation period for Omicron (reportedly an average of 3 days (13) vs. approximately 5-6 days for previous variants) as a halving of the proliferation phase. We also investigate the “unvaccinated” viral load curve from Kissler et al. (longer clearance phase), and the combination of halved proliferation phase and vaccination status.

**Figure 1:**
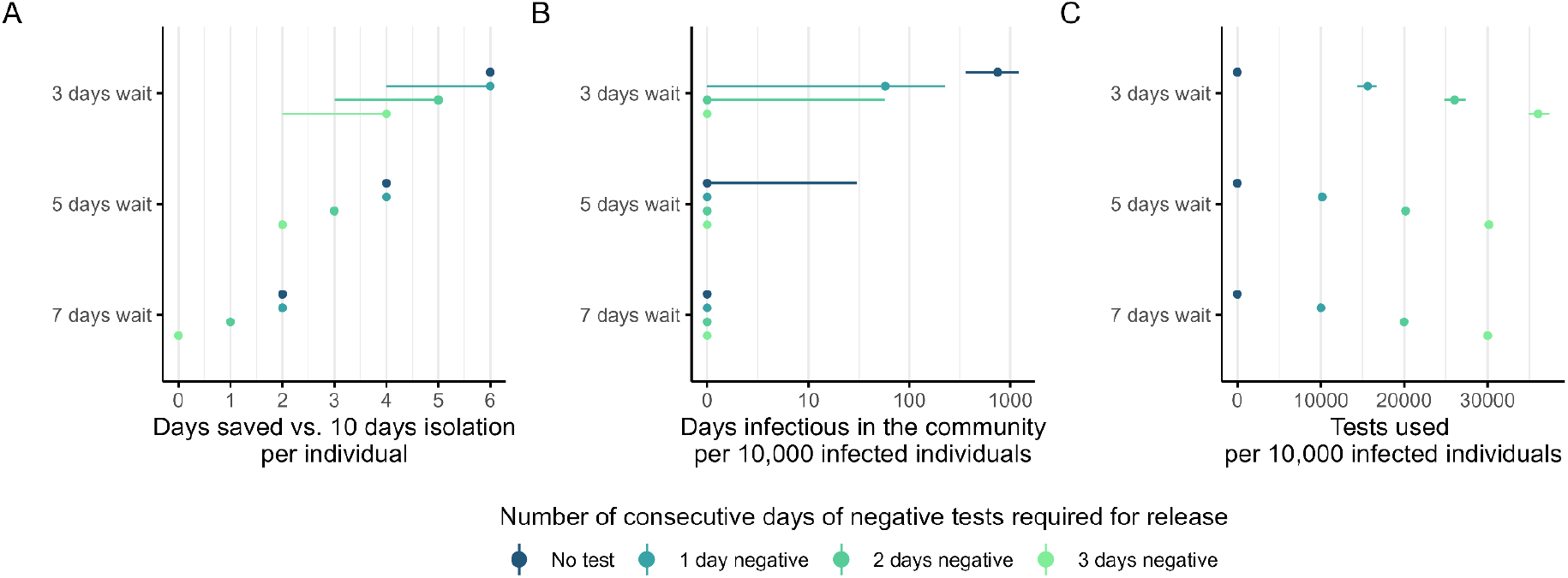
Comparison of policy outcomes for vaccinated populations. A) Number of days saved vs. a 10-day isolation policy per individual and B) days infectious in the community per 10,000 infected individuals following release from isolation for 3, 5, and 7 days wait after an initial positive test to initiate testing and number of consecutive days of negative tests required for release. Points indicate median and error bars represent the 95% uncertainty interval. For days saved, the first day testing positive is considered the minimum mandatory isolation, so for example, a “3 day wait” is equal to 1 day + 3 days wait to then test again on day 4.

## Results

Using an individual-based model of viral load trajectories, we estimate that any policy option requiring individuals to test negative for at least 2 consecutive days is likely to minimise the number of days spent infectious in the community. Minimal days of infectiousness can be achieved with waits as short as 3 days before resuming testing, which maximises the days saved vs. a 10 day isolation period but requires a greater number of tests, as individuals will test for a longer duration before testing negative consecutively. If testing is scarce, or completely unavailable, then 5 or 7 days following a positive test also results in minimal additional transmission, at the expense of longer isolation periods.

For unvaccinated populations, test-to-release policies save fewer days vs. a 10 day isolation, more infectious days are expected, and a greater number of tests will be required as a result of a longer clearance phase and overall duration of infection and test positivity compared to vaccinated populations (Figure S1, Figure S4). Assuming Omicron has a 50% shorter duration of proliferation in both vaccinated and unvaccinated individuals (i.e, faster viral replication and time to infectiousness) increases the number of days saved, reduces the infectious days expected and decreases the number of tests required as individuals are infectious for a shorter period compared to individuals infected with other variants (Figure S1, Figure S4).

## Discussion

Given the likely incidence of Omicron infection in key worker populations, standard 10-day fixed-length self-isolation policies may lead to excess work force absences or substantial under-compliance with testing to avoid absences. As such, some countries such as the UK are revising their isolation policies (14). Our analysis, albeit with pre-Omicron parameters, indicates very low risk associated with reducing the fixed isolation period to 7 days which may be even further reduced when combined with test-to-release schemes such as a 3 or 5 day wait followed by daily testing until receiving 2 consecutive days of negative results. For professions facing critical staffing shortages, shorter isolations with fewer days of negative tests to release may be considered with a small increase in risk.

This work has several limitations. We use a viral load model based on pre-Omicron parameters (10) due to a lack of data on the viral kinetics in Omicron infections. Evidence from pseudovirus infectivity assays (15) and early estimates of a shorter incubation period (13) may indicate faster proliferation of Omicron, for which we conduct a sensitivity analysis assuming a 50% shorter proliferation period (Figure S4). In this analysis, the shortened proliferation phase in effect results in faster relative time from exposure to clearance. However, whether the peak to clearance phase is lengthened is yet unknown, as is the potential interaction with vaccination (in particular boosters), which also reduces the duration of clearance (10), or prior infection. Also unknown is whether peak viral loads are higher or lower in Omicron-infected individuals and the sensitivity of different LFTs to Omicron given changes to the Nucleocapsid (N) antigen, though early assessment indicates sensitivity of tests licensed in the UK is similar to previous variants (8). We model infectiousness as equal to the probability of culturing virus for a given viral load (11) though a like-for-like comparison indicates LFTs to be more sensitive than culture (Figure S1), which may be a result of the difficulty and limits of detection when culturing live virus or alternatively a short period of residual LFT positivity after the infectious period (16). If individuals test positive for over 10 days then it may be prudent to truncate the isolation period regardless of test status, though this may be associated with a small increased risk of transmission. We consider only those isolating after a positive test when testing daily and not the effect of isolating from symptom onset, for which specificity and the timing relative to detectability and infectivity may vary by variant and vaccination status, or for individuals who may test positive when testing less frequently. If individuals are later in their infectious period when developing symptoms or initially testing positive, this may allow for even shorter isolation periods. Individuals should seek a test and isolate upon developing any SARS-CoV-2 symptoms in order to reduce the risk of transmission early in the infectious period. Data from daily sampling studies of Omicron-infected individuals will be able to better inform our assumptions.

Based on a model of viral load kinetics, infectivity, and test sensitivity, shortening the isolation period for SARS-CoV-2 test positive individuals carries minimal risk which can be reduced further by requiring consecutive negative lateral flow tests to release.

## Data Availability

The code and data to reproduce the analysis can be found at https://www.github.com/bquilty25/daily_testing

## Code and data availability

The model was coded in R version 4.1.2 (17). The code to reproduce the analysis can be found at https://www.github.com/bquilty25/daily_testing

## Acknowledgements

We thank Suzanne Pickering, Rui Pedro Galão and Stuart J. D. Neil for sharing raw data on culture positivity and LFT detection from their study (11), and Samuel Clifford and W John Edmunds for their prompt review of the manuscript.

## Supplementary appendix

**Figure S1:**
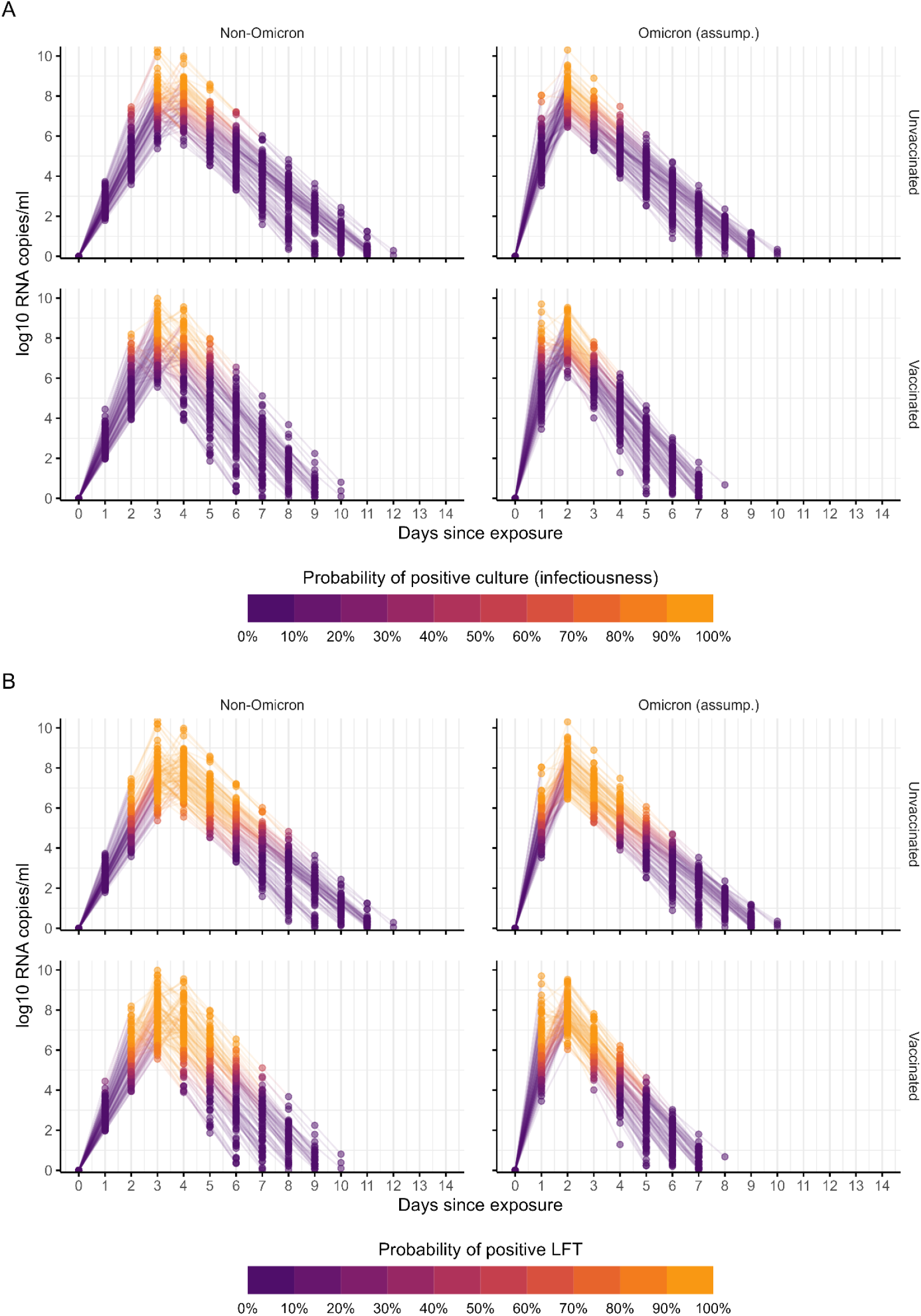
Modelled viral load trajectories and daily probabilities from Kisser et al. (10) with the A) probability of culturing virus and B) testing positive by the Biotime Innova LFT given daily viral load from Pickering et al. (11).

**Figure S2:**
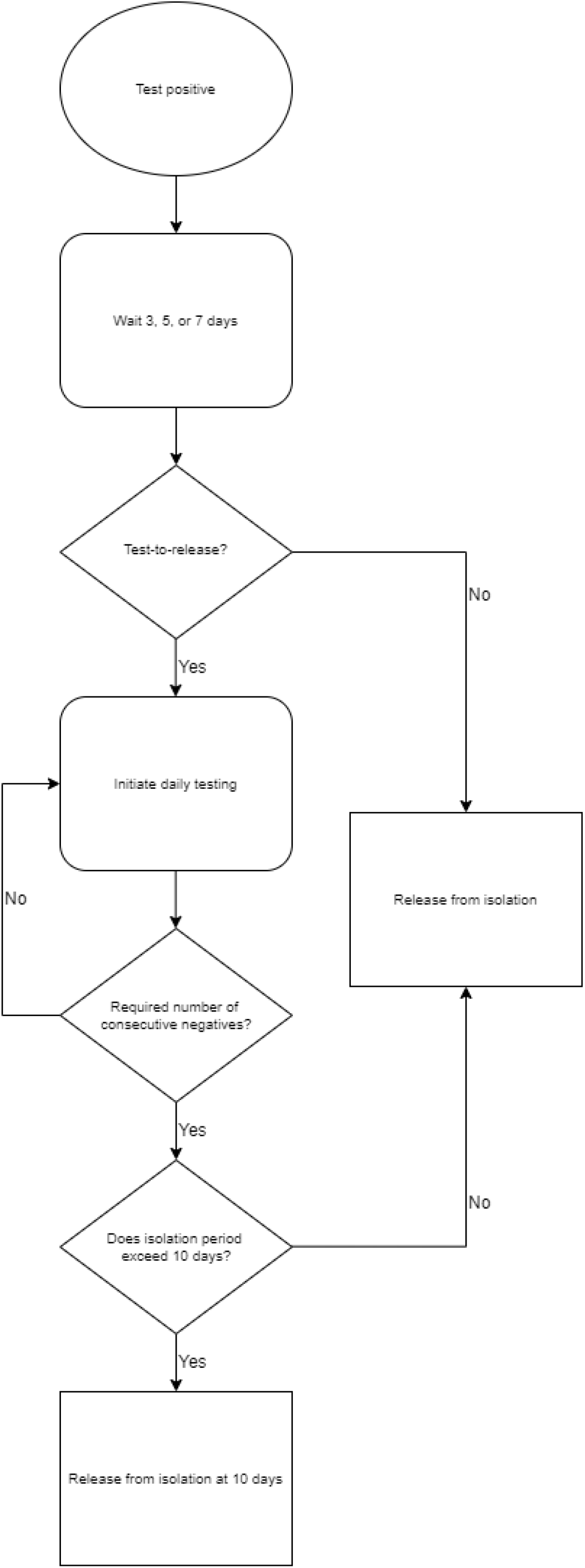
Modelled isolation and testing trajectories.

**Figure S3:**
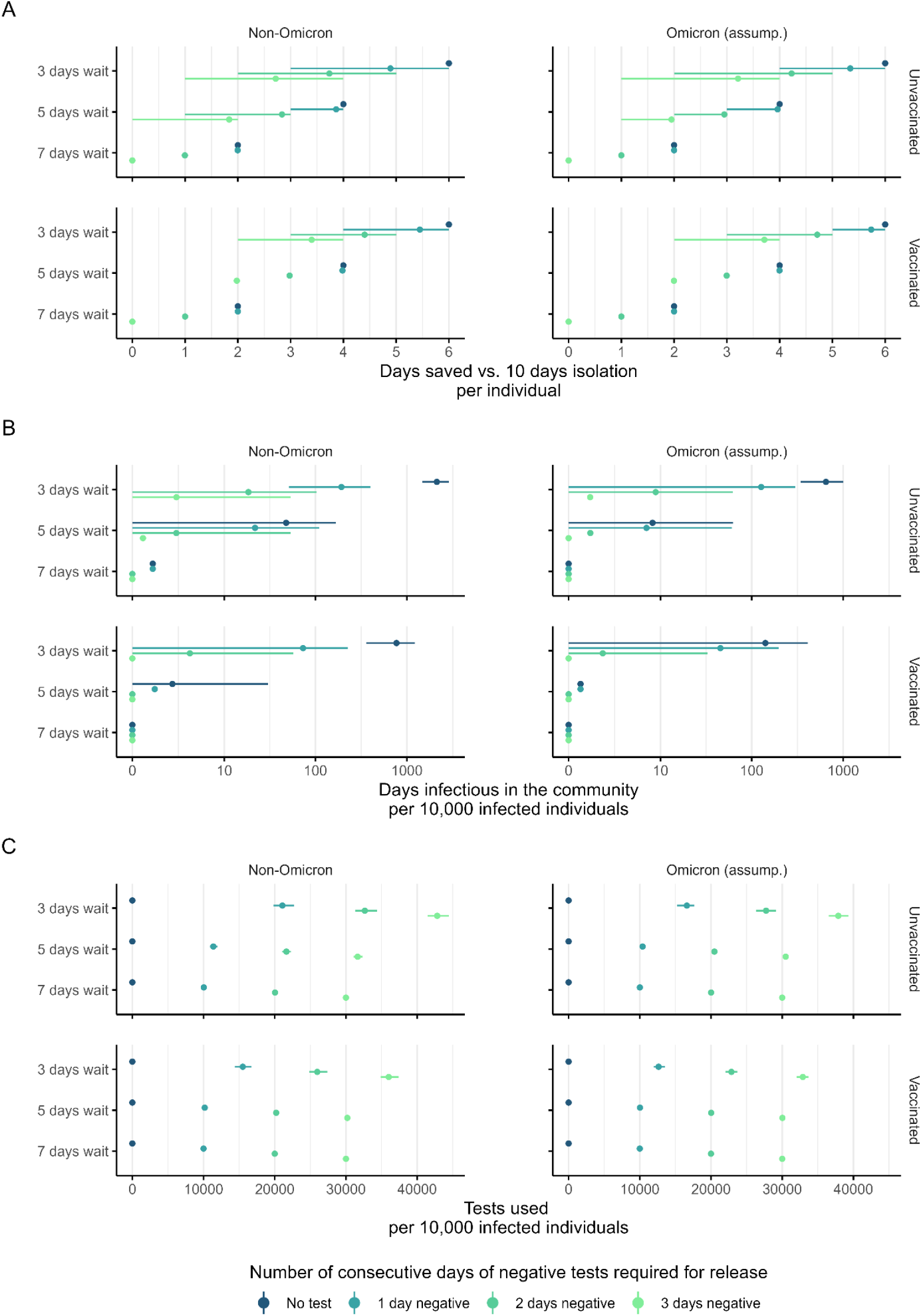
A) Number of days saved vs. 10 days isolation and B) days infectious in the community per 10,000 infected individuals for 3, 5, and 7 days wait after an initial positive test to resume testing and number of consecutive days of negative tests required for release. Points indicate mean and error bars represent 95% uncertainty interval. Viral load trajectories from Kissler et al. (10), with assumed Omicron viral load trajectories equal to non-Omicron viral load trajectories but with a 50% shorter proliferation phase. For days saved, the first day testing positive is considered the minimum mandatory isolation, so for example, a “3 day wait” is equal to 1 day + 3 days wait to then test again on day 4.

